# Molecular methods enhance the detection of pyoderma-related *Streptococcus pyogenes* and *emm*-type distribution in children

**DOI:** 10.1101/2024.02.15.24302883

**Authors:** Jennifer N. Hall, Edwin P. Armitage, Elina Senghore, Saffiatou Darboe, Momodou Barry, Janko Camara, Sulayman Bah, Alexander J. Keeley, James S. McCarthy, Pierre Smeesters, Claire E. Turner, Thomas C. Darton, Michael Marks, Adrienn Angyal, Thushan I. de Silva

**Author notes:** ***Corresponding author:*** Jennifer Hall, Division of Clinical Medicine, School of Medicine and Population Health, University of Sheffield, Sheffield, United Kingdom.

## Abstract

**Background:** *Streptococcus pyogenes*-related skin infections are increasingly implicated in the development of rheumatic heart disease (RHD) in lower-resourced settings, where they are often associated with scabies. The true prevalence of *S. pyogenes*-related pyoderma may be underestimated by bacterial culture.

**Methods:** A multiplex qPCR for *S. pyogenes, Staphylococcus aureus* and *Sarcoptes scabiei* was applied to 250 pyoderma swabs from a cross-sectional study of children <5 years in The Gambia. Direct PCR-based *emm*-typing was used to supplement previous whole genome sequencing (WGS) of cultured isolates.

**Results:** Pyoderma lesions with *S. pyogenes* increased from 51% (127/250) using culture to 80% (199/250) with qPCR. Compared to qPCR, the sensitivity of culture was 95.4% for *S. pyogenes* (95% CI 77.2-99.9) in samples with *S. pyogenes* alone (22/250, 9%), but 59.9% (95% CI 52.3-67.2) for samples with *S. aureus* co-infection (177/250, 71%). Direct PCR-based *emm*-typing was successful in 50% (46/92) of cases, identifying 27 *emm*-types, including six not identified by WGS (total 52 *emm*-types).

**Conclusions:** Bacterial culture significantly underestimates the burden of *S. pyogenes* in pyoderma, particularly when co-infected with *S. aureus*. Molecular methods should be used to enhance the detection of *S. pyogenes* in surveillance studies and clinical trials of preventative measures in RHD-endemic settings.

## Introduction

Superficial bacterial skin infections (pyoderma) are common in childhood and ∼162 million children globally are affected at any one time [1]. A high prevalence is seen in low- and middle-income countries, and in marginalised groups within high-income countries such as indigenous populations [1–3]. *Streptococcus pyogenes* and *Staphylococcus aureus* are the dominant causative pathogens, although microbiological data from pyoderma studies conducted in these settings are scarce [1]. Previous studies have shown that polymerase chain reaction (PCR) improves the detection of *S. pyogenes* in pharyngitis, but the use of molecular assays for *S. pyogenes* in cases of pyoderma has been infrequently evaluated [4–6]. In addition to a broad range of acute infections, post-infectious immune-mediated sequelae of *S. pyogenes* include acute post-streptococcal glomerulonephritis and acute rheumatic fever (ARF), leading to rheumatic heart disease (RHD) [7]. RHD following *S. pyogenes* pharyngitis is well-described, but there is growing recognition of the role of *S. pyogenes* pyoderma in driving RHD in settings with the highest burden of disease [8–10].

Pyoderma caused by both *S. pyogenes* and *S. aureus* is also frequently associated with *Sarcoptes scabiei* infection (scabies) in RHD-endemic settings, which in turn is independently associated with poverty and overcrowding [11–14]. The recent development of a PCR assay to detect *S. scabiei* offers potential for a non-invasive, objective diagnosis of scabies infection and integration alongside diagnostics for pyoderma [15].

Strain typing is an important component of epidemiological surveillance of *S. pyogenes* infections, commonly carried out by sequencing the *emm* gene that encodes the M surface protein [16]. The M protein has also been identified as a major *S. pyogenes* vaccine target, therefore characterisation of *emm*-type distribution is essential to ensure adequate vaccine coverage globally [17]. In high-income settings, the majority of *S. pyogenes* infections are attributed to a small number of different *emm*-types, however, much greater genetic diversity exists in resource-poor settings, with no clear *emm-*type dominance [18].

We previously conducted a cross-sectional study of 1441 children in The Gambia which identified pyoderma in 250/1441 (17%) of children under five years old (95% CI 10–28). Wound swab cultures yielded *S. aureus* in 81% and *S. pyogenes* in 51% of cases. Scabies infection (diagnosed clinically) was seen in 16% of children (95% CI 12–20) and was significantly associated with pyoderma [19]. Whole genome sequencing (WGS) of cultured samples demonstrated diverse *emm*-types [20]. Here, we use a newly-established multiplex quantitative PCR (qPCR) assay for *S. pyogenes*, *S. aureus* and *S. scabiei*, applied to wound swab samples from this study, to explore whether the presence of these pathogens was underestimated by clinical criteria and bacterial culture. Furthermore, we attempt to enhance our data on strain diversity by using direct PCR-based *emm-*typing on *S. pyogenes* qPCR-positive samples that were either culture negative, or where WGS of cultured isolates failed.

## Methods

### Study design, sampling and bacterial culture

Samples were taken during a cross-sectional, cluster-randomised, population-based study in Sukuta, a peri-urban region in the Gambia, as previously described [19]. Ethical approval was provided by the Gambia Government/MRC Joint Ethics Committee (SCC1587). Pyoderma was defined as any skin lesion with evidence of pus or crusts. Children <5 years old within all households in each cluster underwent skin examination by trained research nurses. Skin lesions were clinically classified as scabies, infected scabies, pyoderma, or fungal skin infection. Where pyoderma was diagnosed, superficial saline cleansing was done followed by a single Nylon flocked swab (Copan) collected from the largest lesion into liquid Amie’s transport medium. Swabs were removed from transport media and inoculated the same day on 5% sheep’s blood agar and incubated overnight at 37°C. *S. aureus* and *S. pyogenes* were identified through purity plates (where there was mixed infection), catalase, and agglutination testing (Remel Staphaurex Plus or Streptex latex, Thermo Fisher Scientific). Transport media were stored at -80°C until DNA extraction.

### DNA extraction and qPCR

DNA was extracted from 500μL of transport media using the QIAmp DNA Mini Kit (Qiagen). A pellet formed by centrifugation at 7000 xg for 5 minutes was resuspended in 180μL enzymatic lysis buffer containing 40μL each of lysostaphin (1mg/ml) and lysozyme (100mg/ml). An incubation at 37°C for 45 minutes was followed by addition of 25μL proteinase K and 200μL AL buffer before a further incubation at 56°C for 60 minutes. DNA purification was then carried out according to manufacturer instructions.

Bacterial loads were quantified using standard curves generated by eight 10-fold serial dilutions of extracted DNA from *S. pyogenes* reference strain H293, *S. aureus* strain SH1000 and linearised plasmid DNA containing the *S. scabiei* SSR5 microsatellite sequence [15,21]. Dilutions ranged from 10,000,000 to 1 genome copy per PCR reaction.

Previously described *S. pyogenes* (*speB* gene), *S. aureus* (*nuc* gene), and *S. scabiei* (SSR5 microsatellite) PCR assays were integrated to establish a multiplex qPCR (Luna Universal Probe qPCR Master Mix (New England Biolabs) and primers and probes as outlined in **Supplementary table 1**) [15,21–23]. Nuclease-free water replicates were included as PCR negative controls. Thermocycling conditions consisted of an initial ten minutes at 95°C, followed by 40 amplification cycles of 94°C for 15 seconds and 58°C for 40 seconds. Limits of detection (LOD) were determined using standard curves generated by eight 2.5-fold serial dilutions from 1000 to 1.64 genome copies per PCR reaction, run in 11 replicates. The LOD was defined as the lowest genome copy number that was amplified at a 95% detection rate [24]. When tested in two replicates, the LOD was 31.1 copies for *S. pyogenes,* 3 copies for *S. aureus*, and 4.5 copies for *S. scabiei*. Samples were run in duplicate, with the assay repeated if high intra-replicate variation was seen (cycle threshold (Ct) standard deviation greater than 0.5). Amplification curves were reviewed to ensure consistency with true target amplification. Samples were deemed positive if there was target amplification and a copy number above the LOD.

### Direct PCR-based emm-typing

The *emm* gene was amplified from swab DNA extracts using US Centers for Disease Control and Prevention primers CDC1 (5ʹ-TATTSGCTTAGAAAATTAA-3ʹ) and CDC2 (5ʹ-GCAAGTTCTTCAGCTTGTTT-3ʹ) [25]. PCR was performed with GoTaq polymerase (Promega) as per manufacturer’s instructions, with each reaction containing 200nM of the primers and 10μl of DNA extract. Reaction conditions were at 94°C for 1 minute; followed by 30 cycles of 94°C for 15s, 47°C for 30 seconds, 72°C for 85 seconds; and final extension at 72°C for 7 minutes. PCR products were cleaned (Monarch PCR and DNA Cleanup Kit, New England Biolabs) and eluted into 10μl, then used as the template for nested PCR using the same conditions and primers CDC1 and CDC3 (5’-TTCTTCAAGCTCTTTGTT-3’). After gel visualisation, bands corresponding to the *emm* gene (∼1100bp) were purified (Monarch DNA Gel Extraction Kit, New England Biolabs) and sent for Sanger sequencing (Genewiz, Azenta). If no band was seen on gel visualisation, the first two PCR rounds were repeated from genomic DNA, followed by a 3^rd^ round nested PCR using high-fidelity polymerase Q5 (New England Biolabs) and primers CDC1 and CDC2 (500nM each). Cycling conditions were 98°C for 30 seconds; followed by 34 cycles at 98°C for 10 seconds, 47°C for 30 seconds, 72°C for 30 seconds; and final elongation at 72°C for 5 minutes.

E*mm*-types were assigned using the CDC *emm*-typing database tool [26]. An 180/180 base match determined an *emm*-subtype; matches less than 180/180 but greater than 90/180 were determined as *emm*-types.

### Data analysis

Statistical analysis was performed using R Statistical Software (v4.2.2; R Core Team 2022). Data were compared using a two-tailed Mann Whitney U test for continuous data or Fisher exact test for categorical data. A p value of <0.05 was considered statistically significant, with the Benjamini-Hochberg procedure applied for multiple hypothesis testing. Raw p values are reported where they remain statistically significant after adjusting for multiple testing; others are reported as not statistically significant. The Simpson Reciprocal Index of diversity was calculated for samples with *emm-*types defined by WGS alone, and following addition of direct-PCR typed samples [27,28].

## Results

### Quantitative PCR shows greater diagnostic yield than bacteriological culture

All transport media samples from pyoderma swabs underwent qPCR (n=250). One hundred samples (40%) had either *S. pyogenes* or *S. aureus* detected by qPCR but not culture (**Supplementary table 2**). The greatest additional diagnostic yield was in *S. pyogenes,* identified in 72/250 samples (29%) by qPCR but not culture, resulting in 80% of pyoderma cases being positive for *S. pyogenes*. All *S. pyogenes* culture-positive samples were also qPCR-positive, but 8/250 samples (3%) that were *S. aureus* culture-positive were *S. aureus* qPCR-negative. *S. aureus* and *S. pyogenes* co-infection was seen in 71% (177/250) of samples by qPCR compared to 42% (104/250) by culture (**Supplementary figure 1**). Compared to qPCR, the sensitivity of culture was 95.4% for *S. pyogenes* (95% CI 87-100) and 91.1% for *S. aureus* (95% CI 83-99) in samples in which a single bacterial pathogen was identified, but 59.9% for *S. pyogenes* (95% CI 53-67) and 86.4% for *S. aureus* (95% CI 81-91) from samples in which co-infection was present.

For both *S. pyogenes* and *S. aureus,* qPCR bacterial load was significantly higher in samples that were culture positive compared to those that were culture negative (both p<0.0001) (**Figure 1A and 1B**). *S. pyogenes* load in co-infected samples was significantly lower than in *S. pyogenes* mono-infections (p=0.00094), whereas *S. aureus* load was higher in co-infected samples than in those with *S. aureus* alone (p=0.00078, **Figure 1C and 1D**).

**Figure 1.**
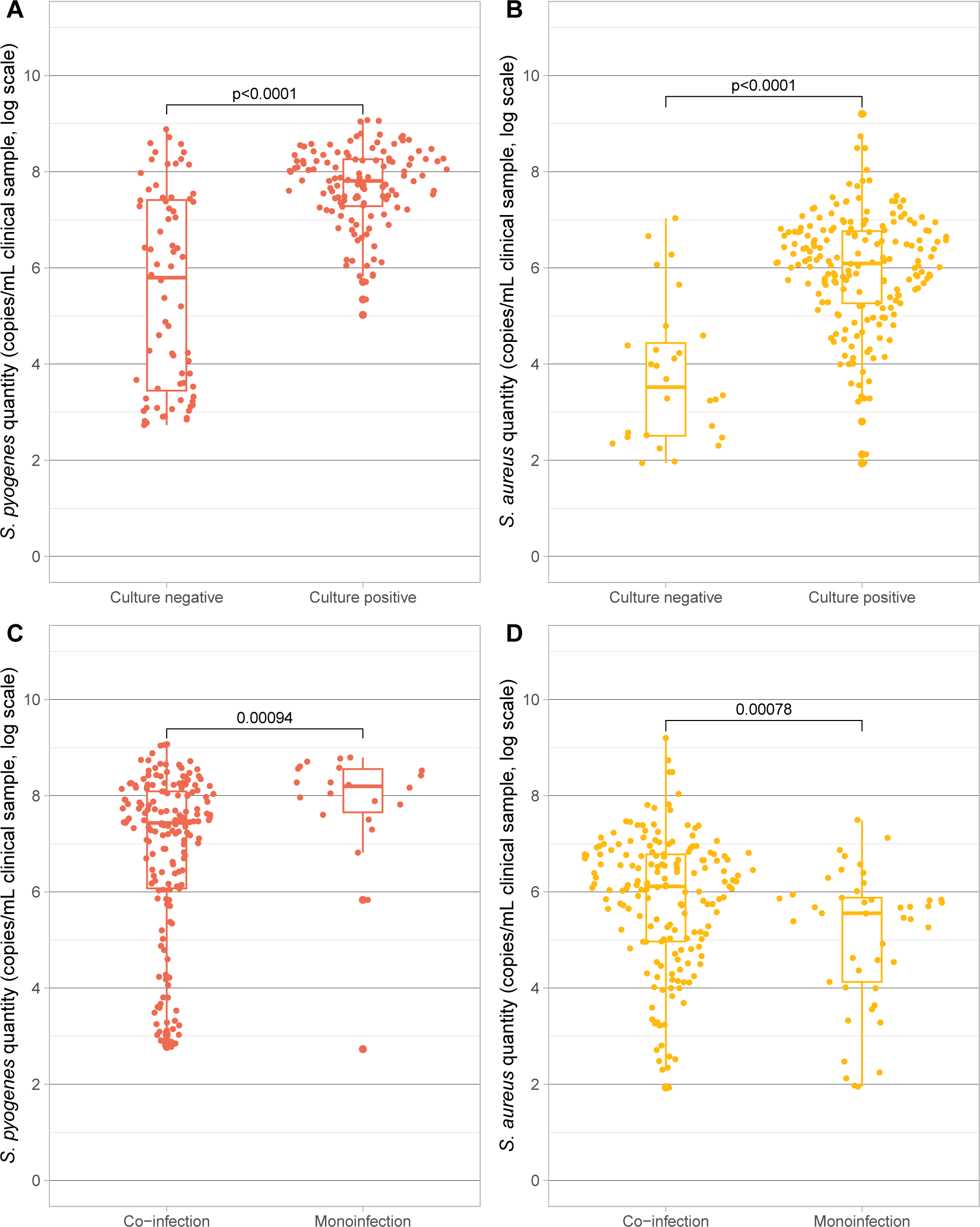
Bacterial quantity in PCR-positive samples for (**A**) S. pyogenes and (**B**) S. aureus by bacteriological culture status; and for (**C**) S. pyogenes and (**D**) S. aureus in co-infected samples compared to samples with a single identified bacterial pathogen. Statistical differences determined using a two-tailed Mann Whitney U test.

Forty three of 250 pyoderma swabs (17%) tested positive by qPCR for *S. scabiei*. Of the 25 swabs from lesions classified clinically as infected scabies, only four (16%) were positive by qPCR (**Supplementary table 3**). In 45 cases where scabies was diagnosed clinically in another body site, seven (16%) were qPCR-positive for *S. scabiei*, along with 18% (32/180) samples from pyoderma cases where no clinical scabies diagnosis was made.

### Bacterial aetiology and load associations with age, body site and season

The proportion of individuals with each pathogen increased with age (**Figure 2**). Within those with pyoderma, age category was significantly associated with pattern of infection as defined by qPCR result (Fisher exact test, p = 0.039), with no participants <1 year having infection with *S. pyogenes* alone, and fewer participants having co-infection with both *S. pyogenes* and *S. aureus* compared to older participants. Within all study participants, 3% (10/302) of those under one year of age had pyoderma with *S. pyogenes*, rising to 13% (41/329) in those aged 12-24 months, and 18% (55/300) in those aged 48-59 months. Clinical sample bacterial load of *S. pyogenes* and *S. aureus* did not significantly vary with participant age (**Supplementary figure 2**).

**Figure 2.**
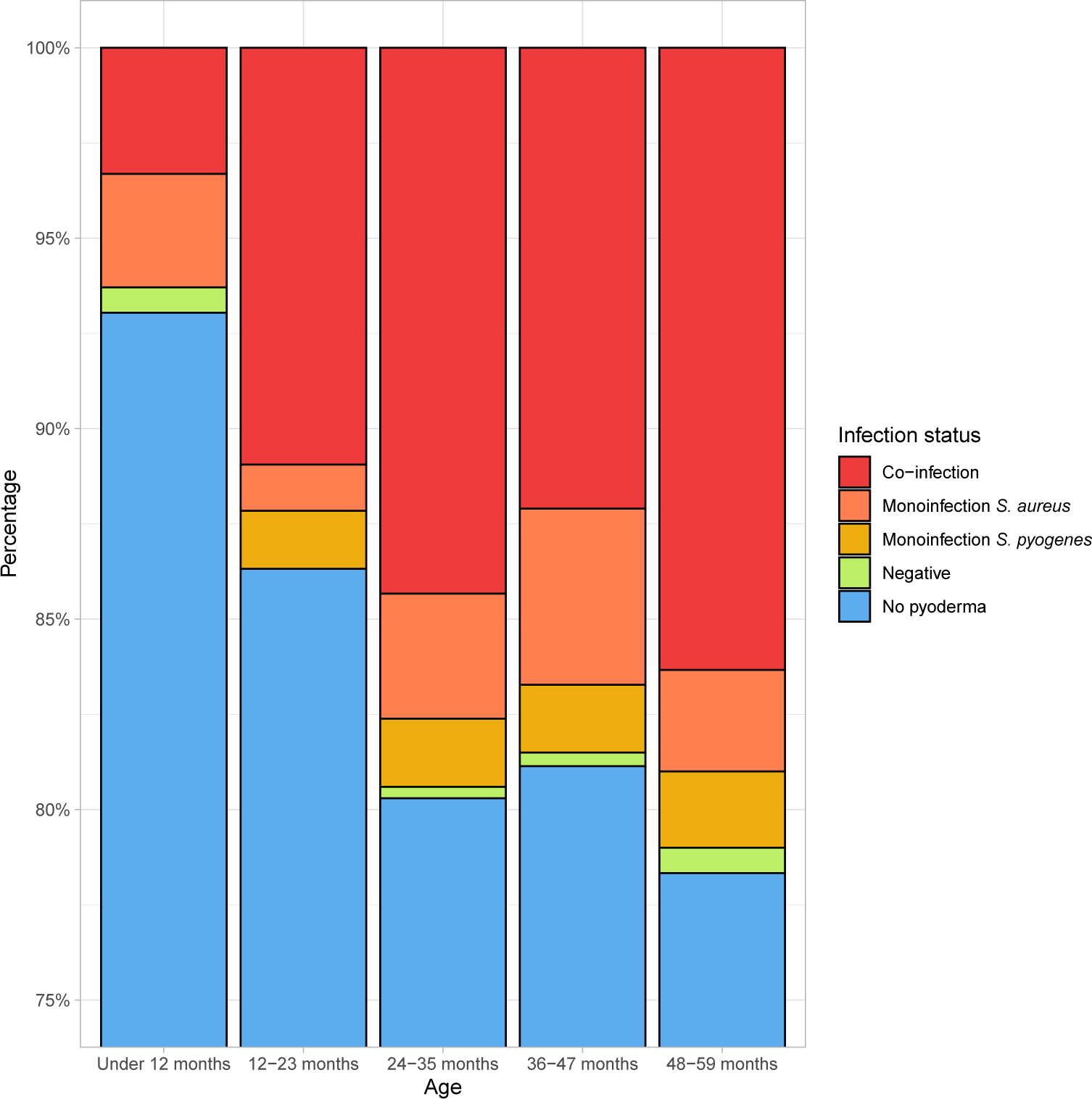
Participant infection status by age, as determined by qPCR result. Those with infection status ‘Negative’ had a clinical diagnosis of pyoderma but neither S. pyogenes nor S. aureus were detected by qPCR.

Pyoderma was most commonly identified above the neck (50% of lesions), followed by leg or foot (33%). Using qPCR, *S. pyogenes* was detected more commonly than *S. aureus* in the lower limb (98% vs 85% of pyoderma lesions; **Supplementary figure 3A**). In contrast, in pyoderma lesions above the neck, *S. aureus* was detected in 91% of lesions compared to *S. pyogenes* in 70%. There was a significant association between site of pyoderma and *S. pyogenes* detection (Fisher exact test, p<0.0001), but not *S. aureus* detection (p = 0.28). A greater *S. pyogenes* load was seen in lower limb compared to above the neck lesions (p<0.0001; **Supplementary figure 3B**), whereas no difference in *S. aureus* load was seen between these sites.

Our previous study reported a significant increase in the prevalence of pyoderma during the rainy season at 23%, compared to 9% before the start of the rains. Co-infections, and mono-infections with both *S. pyogenes* and *S. aureus* increased after the start of the rainy season (**Figure 3A**), but the proportion of each infection type was not significantly different in cases sampled before or after the start of the rainy season (**Figure 3B**). There was, however, a small but statistically significant increase in both *S. pyogenes* and *S. aureus* bacterial loads in pyoderma lesions sampled during the rainy season (**Figure 3C and 3D**).

**Figure 3.**
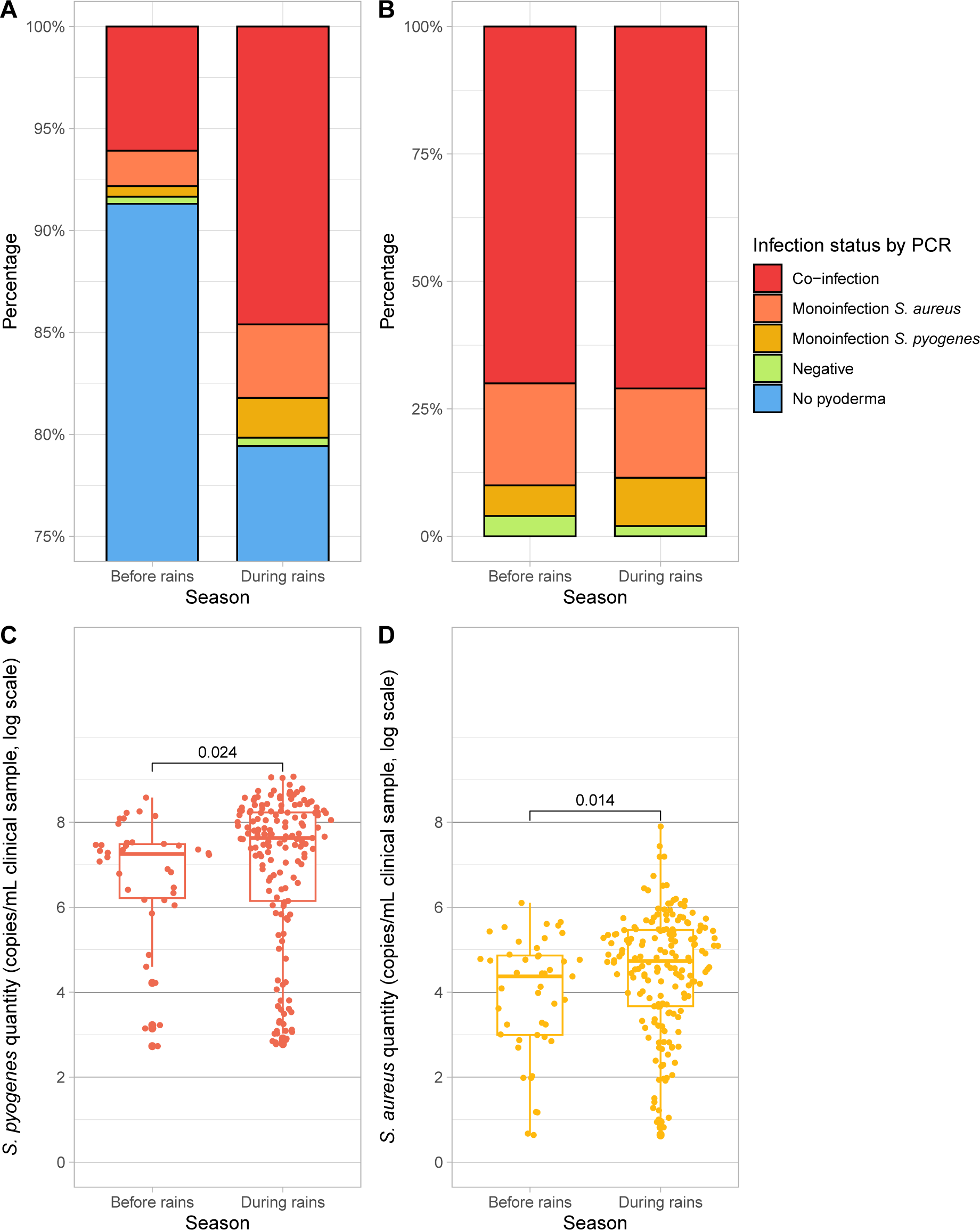
Bacterial aetiology and load of pyoderma cases according to season. Participant infection status by season, as determined by qPCR result, for (**A**) all study participants and (**B**) participants with pyoderma. Those with infection status ‘Negative’ had a clinical diagnosis of pyoderma but neither S. pyogenes nor S. aureus were detected by qPCR, whilst those with status ‘No pyoderma’ did not have swabs taken. Sample bacterial quantity for (**C**) S. pyogenes and (**D**) S. aureus in samples taken before the onset of the rainy season compared to during the rainy season. Statistical differences determined using a two-tailed Mann Whitney U test.

### Emm-type distribution can be enhanced by direct PCR-based typing

Of the samples that were *S. pyogenes* qPCR positive, 127/199 were *S. pyogenes* culture positive and previously underwent WGS. High quality sequence data were obtained for 107/127, from which 46 different *emm*-types were previously reported [20]. *Emm*-typing by PCR was attempted on the remaining 92/199 samples, made up of 20/92 culture-positive samples without high quality WGS data, and 72/92 which were *S. pyogenes* qPCR-positive but culture-negative. *Emm*-types were successfully assigned by direct PCR-based typing for 46/92, with 27 different *emm*-types identified, including six additional *emm-*types not identified by WGS (*emm*66, 73, 82, 102, 111, 208) (**Supplementary table 4**). In *S. pyogenes* qPCR-positive but culture-negative samples, the bacterial load was significantly higher in samples that were successfully typed by direct PCR than untypeable samples (p<0.0001) (**Supplementary figure 4**). The most common *emm*-type identified in the WGS dataset was *emm*80 (6/107), which changed to *emm*4 (10/153) when the direct PCR-typed data were added (**Figure 4A**).

**Figure 4.**
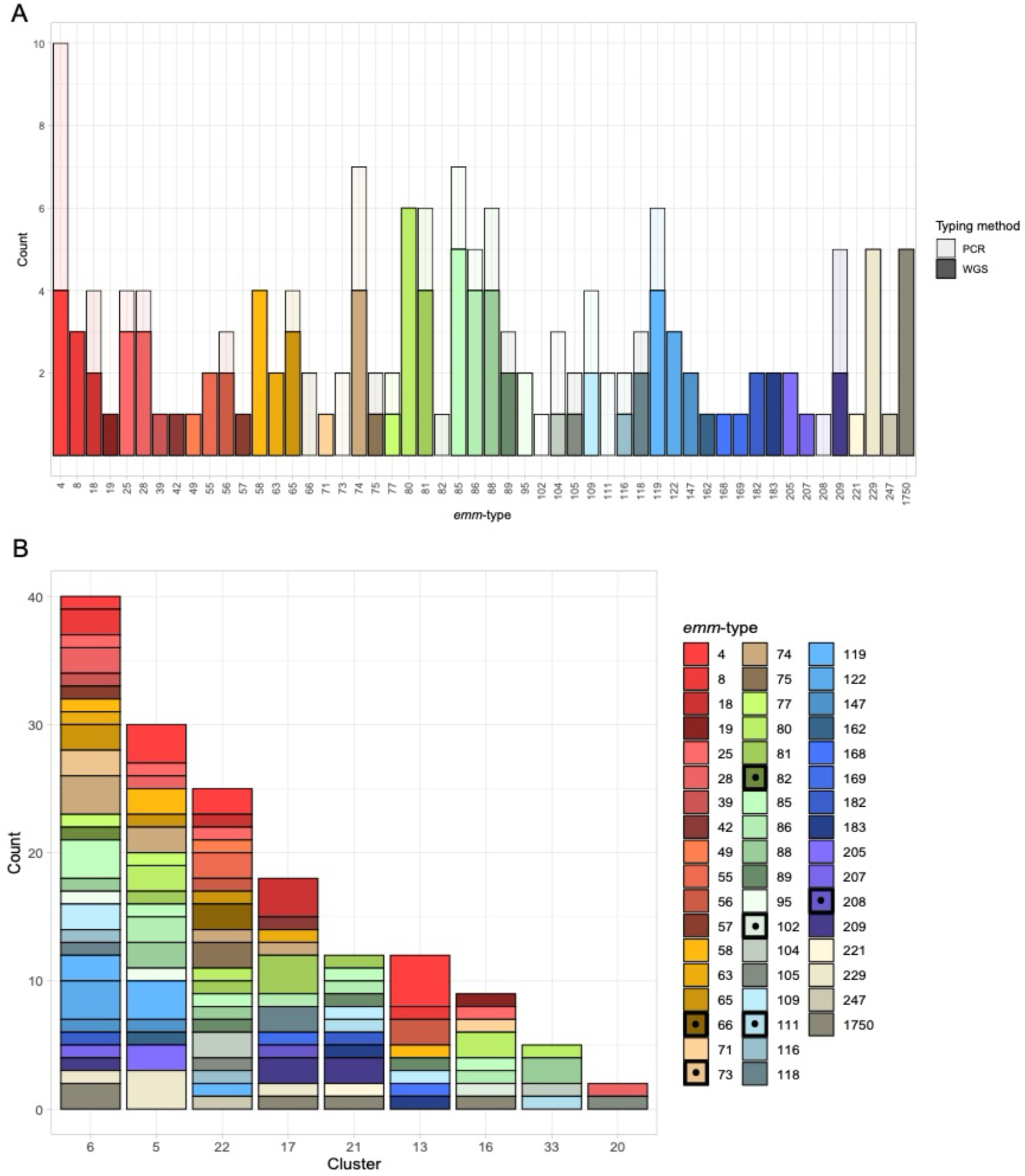
Distribution of S. pyogenes emm-types. Emm-types from 153 of 199 S. pyogenes qPCR positive samples were available. (**A**) Emm-types by typing method. Direct PCR-based emm-typing increased both the number of samples of 21 emm-types in the WGS-defined dataset, as well as detection of six emm-types not in the WGS dataset. (**B**) S. pyogenes emm-type by geographical cluster. Clusters ordered by detected number of pyoderma cases from high to low. Black boxes and dots in the emm-type legend denote emm-types identified by PCR but not WGS.

The Simpson Reciprocal Index of diversity was 49.3 (95% CI 39.0-66.9) for samples *emm-*typed by WGS and 45.6 (95% CI 37.2-59.0) for all *emm-*typed samples. A high degree of *S. pyogenes emm*-type diversity was seen within the nine sampled community geographical clusters (**Figure 4B, Supplementary table 5**). Despite geographical proximity (within and between households), the majority of *emm*-types were represented by a single isolate in a given geographical cluster and limited enrichment for *emm*-types was seen.

## Discussion

In children <5 years old with pyoderma, we demonstrate a significant additional diagnostic yield for *S. pyogenes* using a multiplex quantitative PCR assay in comparison to bacteriological culture alone. Culture-based detection was most impacted when *S. aureus* co-infection was present (sensitivity 59.9%), compared to high sensitivity (95.4%) in *S. pyogenes* mono-infections. Samples with higher bacterial loads were more likely to be culture positive. Co-infections with *S. pyogenes* and *S. aureus* were common (71% of pyoderma lesions), with contrasting impact on bacterial load of the two organisms; lower *S. pyogenes* and higher *S. aureus* load was seen in co-infections compared to mono-infections. The prevalence of both organisms increased significantly with age and bacterial loads increased during the rainy season. We also describe a novel method for *emm*-typing by PCR directly on patient samples without intermediate culture. Direct PCR-based *emm*-typing in *S. pyogenes* qPCR-positive cases where either culture or WGS had failed successfully generated 27 *emm*-types, further enhancing the diverse set of *emm*-types previously generated by WGS.

In studies of pharyngitis, PCR has been demonstrated to have greater yield in the detection of *S. pyogenes* when compared to bacteriological culture [4,5]. Our study shows that culture also underestimates *S. pyogenes* infection in pyoderma lesions, with qPCR identifying *S. pyogenes* in 80% of samples compared to 51% of samples by culture. High rates of co-infection with *S. pyogenes* and *S.aureus* have been described previously, with culture-based studies in First Nations populations in Australia or Canada reporting rates from 29% to 58% [10,29]. We show that most *S. pyogenes* missed by culture are in co-infected samples. Although overgrowth of *S. aureus* during culture and missed *S. pyogenes* colonies may explain this finding, we also find that bacterial loads of *S. pyogenes prior* to culture are lower when *S. aureus* is present. Whether this reflects bacterial inhibition and competition warrants further investigation.

Our original study reported a rise in pyoderma prevalence with age, seen in 7% of those under one year old compared to 21% of those aged three to four years. With more complete detection using qPCR, we demonstrate that *S. aureus* infections are more common at a younger age, with mono- and co-infections with *S. pyogenes* increasing in later childhood. More lower limb lesions were infected with *S. pyogenes* compared to other body sites. This may reflect carriage distribution and transmission patterns, with *S. pyogenes* lower limb pyoderma resulting from behavioural factors and child-to-child transmission. Similarly, *S. aureus* was more common in lesions above the neck, which again could reflect proximity to where *S. aureus* carriage burden may be highest in the nose.

There were considerable differences in our *S. scabiei* qPCR results in comparison to clinical diagnosis, which may be explained by several factors. DNA extraction conditions we used were chosen to enhance extraction from Gram-positive bacteria rather than arthropods [15,30]. Swabs were taken of the pus and crusts associated with pyoderma lesions rather than from skin lesions clinically typical of scabies infection. Most importantly, we compared qPCR result to clinical diagnosis, rather than to a more accurate diagnostic method such as microscopy. Previous studies have highlighted significant variation in clinical diagnoses in both expert and non-expert examiners, and it is possible there are discrepancies between clinical diagnoses and true number of scabies lesions [31,32].

*S. pyogenes* strain diversity was high in our study, with WGS and PCR-based *emm*-typing identifying 52 different *emm*-types across 153 of our isolates. We have previously demonstrated much higher genetic diversity in skin infection isolates from the Gambia compared to the UK [20]. Limited *emm*-type enrichment was seen within geographical clusters, further emphasising the diversity of circulating *emm-*types that are transmitted between children in this setting. This pattern is reflected globally with a higher diversity of *emm*-types seen in low-income settings and may have implications for pathogenesis of RHD, with repeated explosures to different *emm-*types central to auto-immune priming [18,33,34].

We have previously outlined some of the limitations related to study design and sampling in of our original study [19]. The likelihood of bacterial detection and accurate loads are reliant on the quality of the swabbing. Samples had been archived at -80°C for several years prior to undergoing qPCR, with the potential for DNA degradation during that time. A small number of samples were positive for *S. aureus* by culture but not on qPCR, suggesting laboratory contamination of culture plates, misidentification of cultured isolates, or qPCR failure in these samples.

Our findings have implications for the design of interventions against pyoderma, both to reduce the burden of skin disease but also as a constituent of the strategy to address RHD. With the emerging recognition of the role of *S. pyogenes* pyoderma in the development of ARF and RHD, understanding whether low-burden detection of *S. pyogenes* in skin lesions may contribute to the immune priming implicated in RHD pathogenesis will be essential [8–10,34–36]. With restored global interest in the development of a vaccine against *S. pyogenes* and RHD [37], it is critical to have a robust understanding of the epidemiology and strain diversity of *S. pyogenes* infection. Molecular methods should be central in enhanced surveillance for *S. pyogenes* in high-burden settings to aid the design and assessment of future interventions against RHD.

## Data Availability

All data produced in the present study are available upon reasonable request to the authors.

## Footnotes

## Previous presentation

Preliminary data from this study were presented at the British Infection Society Spring Meeting 2023, Manchester, United Kingdom. Data will also be presented at the European Congress of Clinical Microbiology and Infectious Diseases 2024, Barcelona, Spain.

## Corresponding author

Jennifer Hall, Division of Clinical Medicine, School of Medicine and Population Health, University of Sheffield, Sheffield, United Kingdom

