## Supplementary Material for "Molecular methods enhance the detection of pyoderma-related *Streptococcus pyogenes* and *emm*-type distribution in children"

***Supplementary table 1****: Primers and probes used in multiplex qPCR reaction*

| Target | Forward | Sequence | Volume per 25µL reaction (µL) |
|---|---|---|---|
| ***speB*** | Forward primer | 5’ CTAAACCCTTCAGCTCTTGGTACTG 3’ | 0.75 |
| | Reverse primer | 5’ TTGATGCCTACAACAGCACTTTG 3’ | 0.75 |
| | Probe | 5’ Cy5-CGGCGCAGGCGGCTTCAAC-BHQ2 3’ | 0.875 |
| ***nuc*** | Forward primer | 5’ CATCCTAAAAAAGGTGTAGAGA 3’ | 1 |
| | Reverse primer | 5’ TTCAATTTTMTTTGCATTTTCTACCA 3’ | 1 |
| | Probe | 5’ YY-TTTTCGTAAATGCACTTGCTTCAGGACCA-BHQ1 3’ | 0.5 |
| **SSR5** | Forward primer | 5’ GTCGAAGGTGGAGACAAAGATAG 3’ | 1 |
| | Reverse primer | 5’ CGGACTCTTCATCGATTGGTATC 3’ | 1 |
| | Probe | 5’ 6FAM-TCGTTGAGA/ZEN/TGAAGCGACCAGGAGATG-3IABkFQ 3’ | 0.5 |

***Supplementary table 2:*** *Pathogen identification by qPCR compared to bacteriological culture.*

| **Target organism PCR result** | **Target organism culture result** | | **Total** |
|---|---|---|---|
| | **Positive N (%)** | **Negative N (%)** | **N** |
| ***Streptococcus pyogenes*** | | | |
| Positive | 127 (50.8) | 72 (28.8) | 199 |
| Negative | 0 (0) | 51 (20.4) | 51 |
| Total | 127 (50.8) | 123 (49.2) | 250 |
| ***Staphylococcus aureus*** | | | |
| Positive | 194 (77.6) | 28 (11.2) | 222 |
| Negative | 8 (3.2) | 20 (8) | 28 |
| Total | 202 (80.8) | 48 (19.2) | 250 |

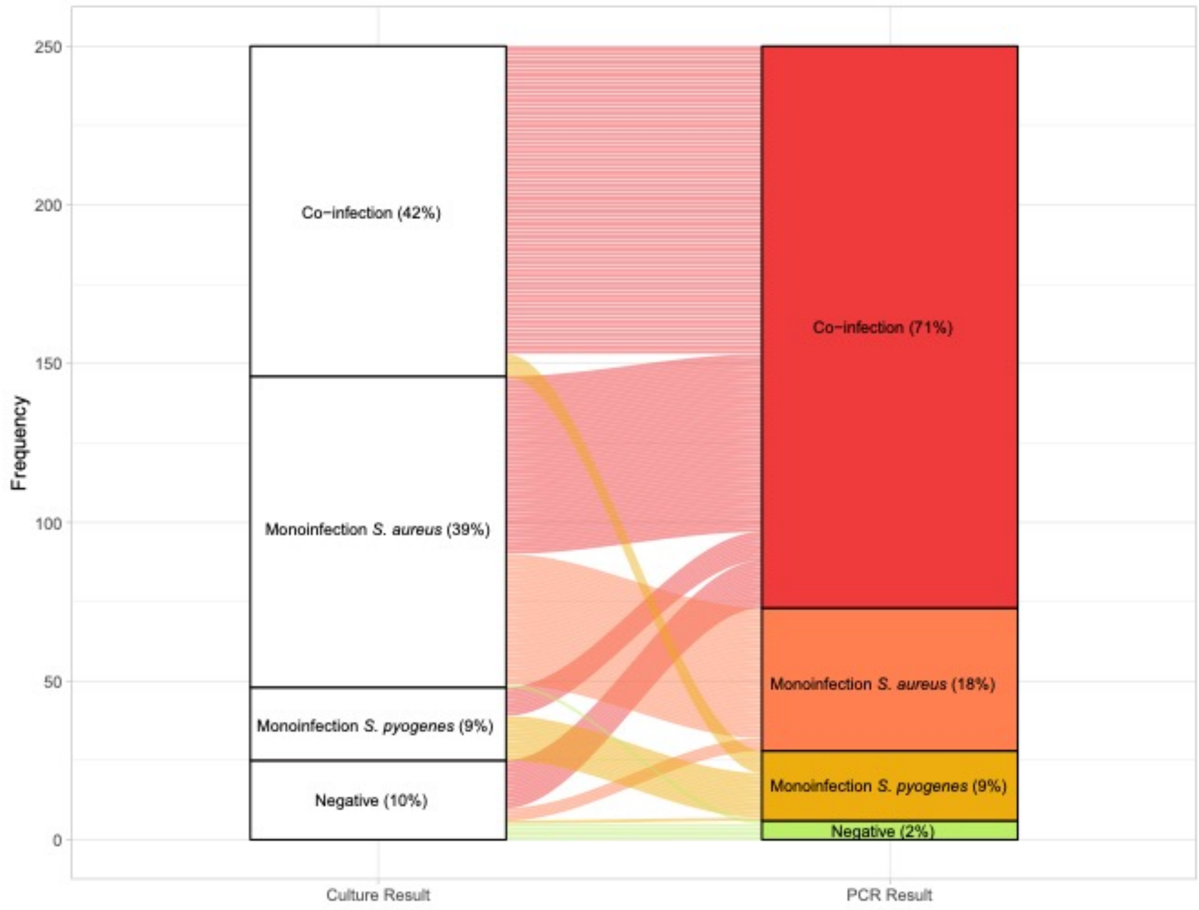


***Supplementary figure 1:*** *Change in distribution of bacteriological results between culture and qPCR.*

***Supplementary table 3****: Identification of S. scabiei by qPCR compared to clinical diagnosis.*

| Clinical diagnosis | PCR positive<br>N (%) | PCR negative<br>N (%) | Total<br>N |
|---|---|---|---|
| Infected scabies | 4 (16) | 21 (84) | 25 |
| Scabies elsewhere | 7 (16) | 38 (84) | 45 |
| No scabies | 32 (18) | 148 (82) | 180 |

*'Scabies elsewhere' = Individuals where scabies and pyoderma were present at different body sites. The swab was taken from the pyoderma lesion.*

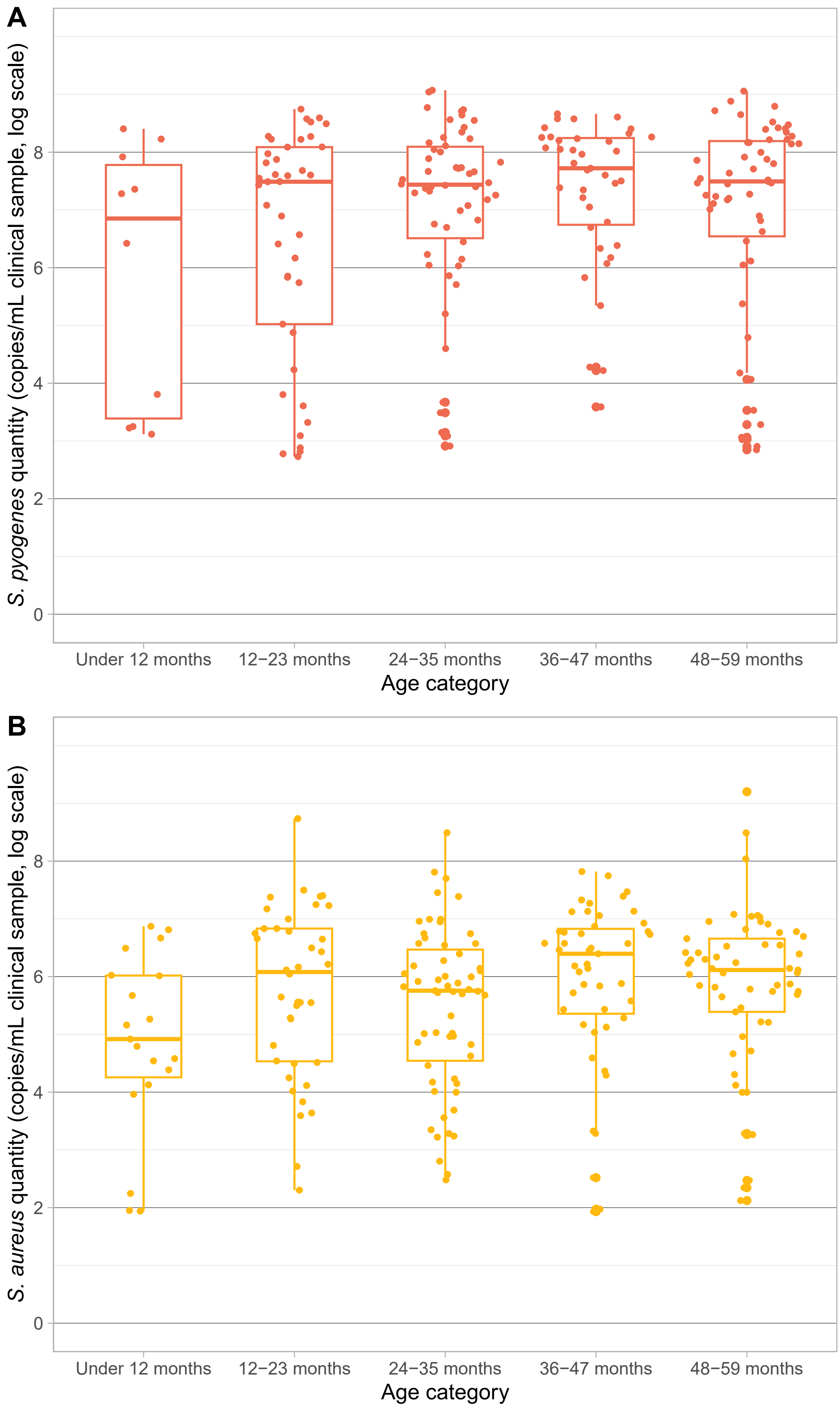


***Supplementary figure 2**: Bacterial quantity in PCR-positive samples for (**A**) S. pyogenes and (**B**) S. aureus, by age category.  Pairwise comparisons using two-tailed Mann Whitney U tests showed no significant difference in quantity between age categories.*

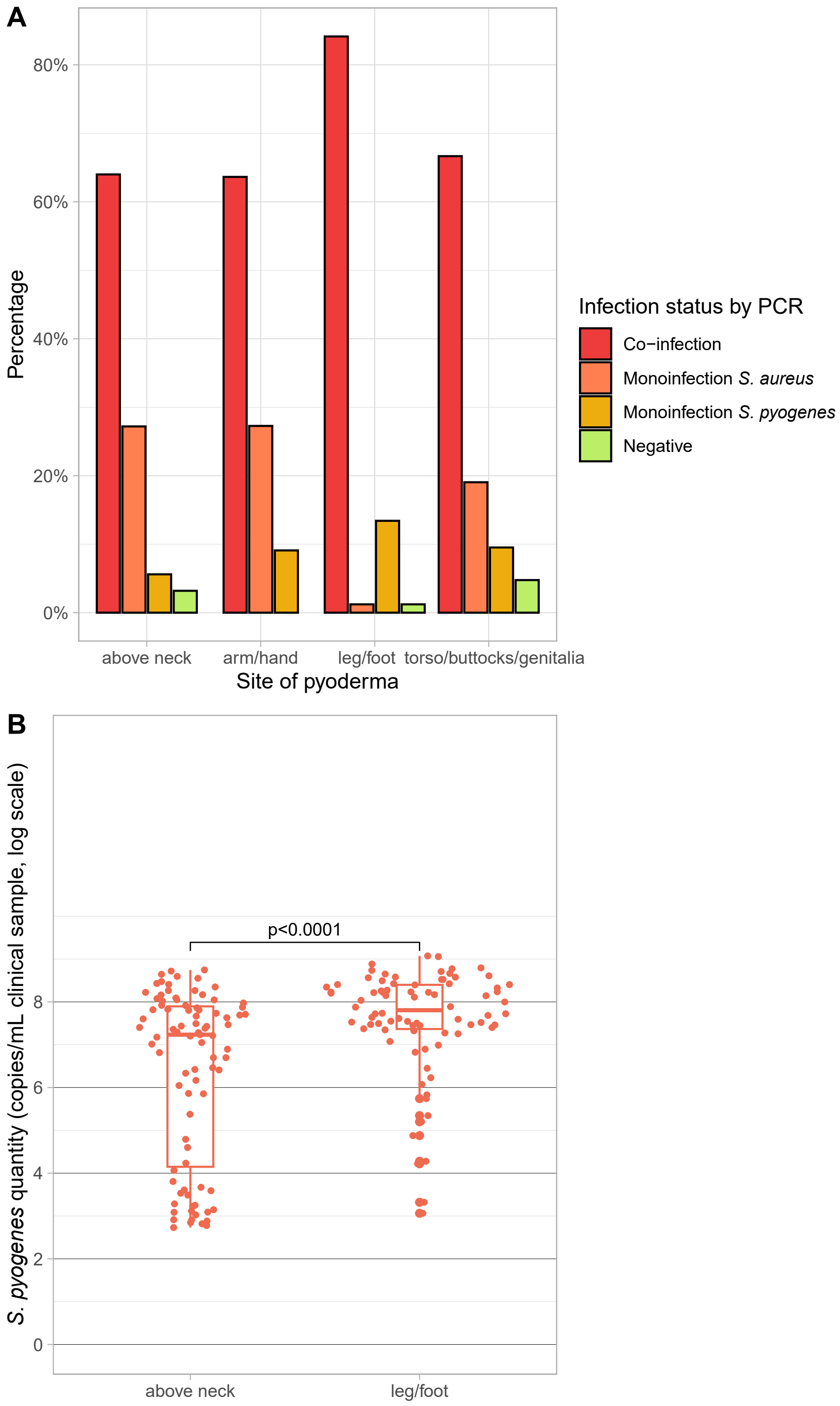


***Supplementary figure 3:*** *(**A**) Participant infection status by site of pyoderma, as determined by qPCR result. Those with infection status 'Negative' had a clinical diagnosis of pyoderma but neither S. pyogenes nor S. aureus were detected by qPCR. (**B**) Sample bacterial quantity for S.*

*pyogenes in samples taken from pyoderma lesions above the neck and in the leg or foot. Statistical differences determined using a two-tailed Mann Whitney U test.*

***Supplementary table 4****: emm-typing by culture status.*

| Typing method | Culture positive, PCR positive N (%) | Culture negative, PCR positive N (%) | Total N |
|---|---|---|---|
| **Emm-typed by WGS** | **107** | **-** | **107** |
| **Emm-typed by PCR** | **19 (41.3)** | **27 (58.7)** | **46** |
| Nested PCR alone | 0 (0) | 27 (100) | 27 |
| Nested PCR + PCR with Q5 | 19 (100) | 0 (0) | 19 |
| **Could not be typed** | **1 (2.2)** | **45 (97.8)** | **46** |

*WGS was attempted on culture-positive samples only.*

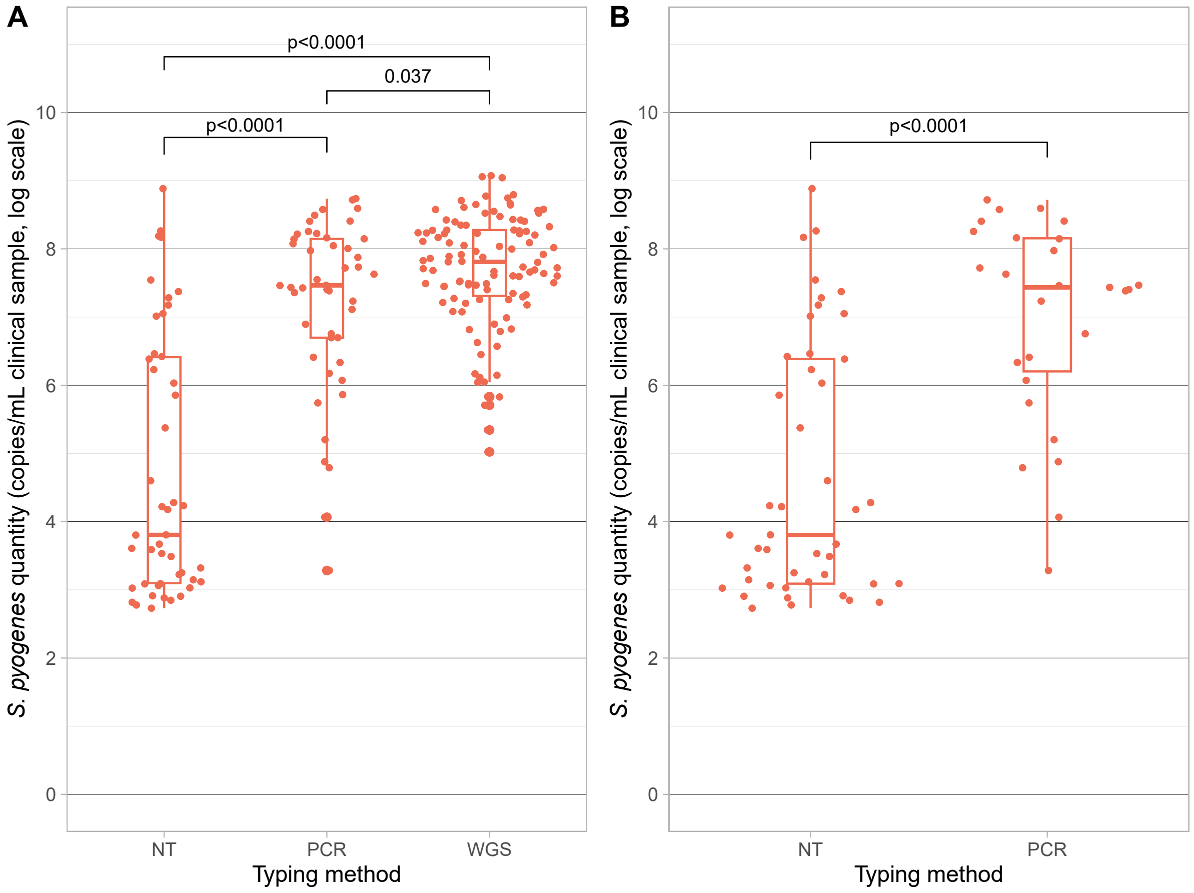


***Supplementary figure 4****: Bacterial quantity in S. pyogenes qPCR-positive samples according to emm-typing method for (A) all samples and (B) culture-negative samples. NT = not typeable*

*by WGS or PCR. Statistical differences determined using a two-tailed Mann Whitney U test. There was no statistical difference in bacterial quantity between samples typed using nested PCR alone compared to samples requiring nested PCR and an additional amplification using Q5.*

***Supplementary table 5:*** *Summary of total participant number, number with pyoderma, number with S. pyogenes on qPCR, and emm-typing, by geographical cluster.*

| **Geographical cluster ID** | **All participants** | **Participants with pyoderma** | ***S. pyogenes* PCR positive** | **No of isolates *emm*-typed (WGS + PCR)** | **No of different *emm*-types** | ***emm*-types (N) identified by PCR but not WGS** |
|---|---|---|---|---|---|---|
| **6** | 277 | 74 | 55 | 40 | 27 | 4 (1); 28 (1); 65 (1); 73 (2); 74 (1); 77 (1); 82 (1); 85 (1); 88 (1); 109 (1); 116 (1); 119 (1); 209 (1) |
| **5** | 82 | 43 | 41 | 30 | 18 | 4 (3); 25 (1); 74 (1); 81 (1) |
| **22** | 68 | 36 | 30 | 25 | 20 | 56 (1); 66 (2); 74 (1); 75 (1); 85 (1); 89 (1); 104 (1); 119 (1) |
| **17** | 306 | 34 | 22 | 18 | 12 | 18 (2); 81 (1); 118 (1); 208 (1); 209 (2) |
| **21** | 243 | 21 | 16 | 12 | 11 | 86 (1); 109 (1); 111 (1) |
| **13** | 113 | 18 | 16 | 12 | 8 | 4 (2) |
| **16** | 189 | 15 | 12 | 9 | 8 | 102 (1) |
| **33** | 207 | 7 | 5 | 5 | 4 | 88 (1); 104 (1); 111 (1) |
| **20** | 62 | 2 | 2 | 2 | 2 | 105 (1) |
| **Total** | **1547** | **250** | **199** | **153** | | |

*Clusters ordered by detected number of pyoderma cases, from high to low.*
